# Impact of air pollution and tobacco smoke on the placental small non-coding RNA landscape

**DOI:** 10.64898/2026.10.16.26365732

**Authors:** Álvaro Falcón-Gracia, Hana Vespalcová, Dolors Peligrí, Sofia Aguilar-Lacasaña, Lorena Pantano, Ramil N. Nurtdinov, Zoraida García, Alan Domínguez, Laura Gómez-Herrera, Olga Sánchez, Mireia Gascón, Maria Foraster, Roderic Guigó, Elisa Llurba, Maria Dolores Gomez-Roig, Ioar Rivas, Payam Dadvand, Jordi Sunyer, Mariona Bustamante, Marta Cosin-Tomas

## Abstract

Fetal development is highly sensitive to environmental influences, including maternal smoking during pregnancy (MSDP) and air pollution. The placenta may play a central role in mediating these effects through epigenetic mechanisms, including small non-coding RNAs (sncRNAs). While most studies have focused on microRNAs (miRNAs), other sncRNA classes remain poorly characterized. Here, we characterized the placental sncRNA landscape and investigated its associations with prenatal exposure to MSDP and air pollution.

The sncRNA expression profile of 390 chorionic villi samples from the Barcelona Life Study Cohort (BiSC) was measured using small RNA sequencing and the seqBuster/seqCluster pipeline. MSDP was assessed via questionnaire, and prenatal exposure to black carbon (BC), particulate matter ≤2.5 μm (PM_2.5)_, and nitrogen dioxide (NO_2)_ was estimated using hybrid dispersion-land-use regression (LUR) spatiotemporal models. Associations were evaluated using linear regressions adjusted for relevant covariates.

We identified 979 placental sncRNA clusters, assigned primarily to miRNAs (33.2%), piRNAs (26.4%), snoRNAs (12.2%), tRFs (7.8%), and others (20.4%). MSDP was associated with the downregulation of miR-421 and the upregulation of multiple snoRNAs from the imprinted *DLK1-DIO3* locus (*SNORD113-4, SNORD114-2, SNORD114-3, and SNORD114-17*), as well as the piR-26394. No sncRNAs remained significantly associated with air pollution after multiple-testing correction, although several miRNAs showed suggestive associations.

Overall, this study provides a comprehensive characterization of placental sncRNAs and identifies previously unreported smoking-associated sncRNAs within the imprinted *DLK1-DIO3* locus, expanding our understanding of placental epigenetic responses to prenatal environmental exposures.

## 1. Introduction

Fetal development is considered a critical window of vulnerability to the influence of environmental factors. According to the Developmental Origins of Health and Disease (DOHaD) hypothesis, exposures spanning the perinatal period can impact disease outcomes in adulthood^1,2^.

Air pollution is a mixture of different pollutants, including particulate matter (PM) and nitrogen dioxide (NO_2)_. Within PMs, those with an aerodynamic diameter ≤2.5 µm (PM_2.5)_, particularly black carbon (BC) and ultrafine particles, are considered especially harmful due to their ability to penetrate deep into the respiratory tract and enter systemic circulation. These exposures along with maternal smoking during pregnancy (MSDP) and other environmental factors, are of particular concern during pregnancy^3^, and various epidemiological studies suggest they can disrupt fetal development and increase the risk of adverse outcomes ranging from pregnancy loss, stillbirth, fetal growth restriction, preterm birth, and congenital anomalies to longer-term effects on offspring neurodevelopment, cardiovascular function, and respiratory health ^4–6^.

The placenta is a transient fetal organ that mediates maternal physiological and environmental signals to the developing fetus and is essential for supporting growth and maintaining pregnancy^7^. It acts as a semi-permeable barrier regulating the bidirectional transfer of a wide range of substances, including gases, nutrients, hormones, metabolites, xenobiotics, and waste products between mother and fetus^8^. After implantation, trophoblast invasion leads to the formation of chorionic villi, the main functional units of the placenta that contribute to endocrine activity, immune regulation, and structural support^9,10^.

Emerging evidence indicates that prenatal exposure to certain air pollutants and tobacco smoke can adversely affect placental structure and function, contributing to pregnancy complications^11^, adverse birth outcomes, and long-term health risks in adulthood^12^, although the underlying mechanisms remain unclear. PM_2.5 h_as been suggested to disrupt placental function by inducing oxidative stress, inflammation, and vascular and angiogenic alterations, while tobacco-related toxins have been associated with impaired placental formation, possibly by reducing uteroplacental blood flow and enhanced inflammatory processes^13^. Nevertheless, the biological mechanisms remain incompletely understood; therefore, placental molecular profiles at birth, including epigenetic and transcriptomic signatures, provide a valuable framework for investigating these pathways^14^.

Small non-coding RNAs (sncRNAs) are short RNA molecules, typically 18-200 nucleotides (nt) long, that act as crucial regulators of diverse cellular processes, ranging from post-transcriptional gene silencing to epigenetic and translational regulation. Among these, microRNAs (miRNAs) are the most abundant and extensively studied species. Their canonical mechanism involves binding to complementary sequences within the 3′ untranslated region (3’UTR) of target mRNAs, though they also interact with 5’UTRs and coding regions. Notably, three specific miRNA clusters located on chromosomes 14 (C14MC) and 19 (C19MC and miR-371-373 cluster)^15^, are predominantly expressed in the placenta and play key roles in trophoblast differentiation, angiogenesis, and maternal-fetal immune tolerance^14,16,17^. Beyond miRNAs, other sncRNA species contribute significantly to regulatory networks. Piwi-interacting RNAs (piRNAs) preserve the germline genome integrity against transposable elements and are critically important in mammalian sperm cells^18^; yet, recent evidence suggests they also exert post-transcriptional and epigenetic regulatory roles in the placenta, modulating genes involved in cell proliferation and invasion^19^. Small nucleolar RNAs (snoRNAs) primarily guide chemical modifications of ribosomal RNAs (rRNAs)^20^; however, they can also be processed into stable snoRNA-derived fragments (sdRNAs), some of which exhibit miRNA-like functions ^21^. Similarly, transfer RNAs (tRNAs) can also be cleaved into tRNA-derived fragments (tRFs) under stress conditions^22^, participating in translation inhibition, retrotransposon silencing, and the regulation of cell survival pathways^23^. Finally, small nuclear RNAs (snRNAs) regulate pre-mRNA splicing as core components of the spliceosome^24^.

Despite its central role in both maternal and fetal health^25^, the placenta remains underrepresented in transcriptomic/epigenomic studies^26^. Moreover, most studies examining placental responses to prenatal environmental exposures have focused on protein-coding genes or miRNAs, leaving the broader landscape of sncRNAs largely unexplored. This gap of knowledge is compounded by analytical frameworks that are predominantly optimized for miRNA analysis, which has limited the characterization of other sncRNA types^26,27^. As a result, only a few studies have profiled placental sncRNAs^19,26,28^, and even fewer have evaluated their relationships with environmental exposures^17,29^.

In this study, we characterized the placental sncRNA profile using a cluster-based bioinformatic pipeline in 390 mother-child pairs from the Barcelona Life Study Cohort (BiSC). We subsequently investigated associations between sncRNA levels and prenatal exposure to air pollution and MSDP.

## 2. Materials and methods

### 2.1. Study population

The study was based on the Barcelona Life Study Cohort (BiSC), an ongoing population-based prospective birth cohort with 1,032 mother-child pairs that aims to understand the effect of early environmental and genetic factors on maternal and child health and development^30^. The details about the recruitment process, follow-ups, and data collection methods are published elsewhere^31^. Briefly, mothers were recruited during their first routine hospital visit around week 12 of their pregnancy at the three major university hospitals in Barcelona: Hospital Sant Joan de Déu (HSjD), Hospital Clínic-Maternitat, and Hospital Sant Pau i la Santa Creu (HSP). The inclusion criteria were: (i) living in the Barcelona metropolitan area, (ii) being between 18 and 45 years old, (iii) having a singleton pregnancy, and (iv) having the intention to deliver in one three recruiting hospitals. Exclusion criteria were (i) residing outside the study area, (ii) being unable to communicate effectively in Spanish, Catalan, or English, and (iii) carrying a fetus with known congenital anomalies. The enrolment of the BiSC participants was conducted between October 2018 and April 2021, and the first data collection was done in the first trimester of pregnancy, with follow-ups in the second and third trimester of pregnancy, delivery, and months 1, 2, 6, 8, 12, 18, 24, and 48 postnatally. This study included 390 mother-child pairs with placental sncRNA data, of whom 359 had complete information on prenatal exposures and covariates.

### 2.2. Placental biopsy, RNA extraction and sncRNA sequencing

Of the 1,032 mother-child pairs followed until birth, 611 placentas were collected. Placenta biopsies of around 2.5 cm (from the maternal to the fetal side) and 1 cm width were obtained from two opposite quadrants at a distance of around 3-4 cm from site of cord insertion. Then, these biopsies were cut in two, giving a total of 4 biopsies of 2.5 x 0.5 cm. Two of them (one from each quadrant) were directly frozen in liquid nitrogen and transferred to -80C for future use. The other two biopsies were treated with RNAlater® solution (Ambion, Inc.) and sent to the laboratory where they were divided in four pieces, corresponding roughly to the fetal membranes, the upper chorionic villi, the lower chorionic villi and the region adjacent to the maternal decidua. Finally, all the biopsies were stored at -80°C for future use.

Before RNA extraction, placental samples were fully randomized. For genomic RNA extraction, a fragment of approximately 30-40 mg from the upper chorionic villi biopsy was dissected in liquid nitrogen. Then, the tissue was disrupted/homogenized using a bead mill (bead beater) at 4°C for 26 seconds. Genomic RNA from the placenta was isolated using the AllPrep®DNA/RNA/miRNA Universal Kit, (Qiagen). RNA quality was evaluated on a NanoDrop spectophotomer (Thermo Scientific) and a 2100 Bioanalyzer Instrument (Agilent) and stored at -80°C until further processing.

From the 611 placentas, 480 RNA samples, including 27 technical duplicates, were selected for sncRNA profiling based on the quality standards: RNA Integrity Number (RIN) ranging from 5.1 to 9.8 with an average of 7.71 (SD = 1.24), and 260/280 ratios close to 2.0.

Quantification of sncRNAs in these 480 placenta RNA samples was conducted through next-generation sequencing platforms at the Genomics Core Facility of the Centre for Genomic Regulation in Barcelona. Samples were randomized before cDNA library preparation, which was performed using the NEBNext Small RNA Library Prep Set for Illumina with 800 ng of RNA input. Size selection (insert: 17-52 nucleotides) was performed via 6% NOVEX polyacrylamide gel electrophoresis. Libraries concentrations were measured by qPCR using the KAPA Library Quantification Kit (KapaBiosystems). Finally, equimolar pools of 40 samples were sequenced on Illumina HiSeq2500 equipment (single end, 50 bp), targeting a minimum of 5 million (M) per sample.

### 2.3. Small non-coding RNA pre-processing

The bioinformatics pipeline was based in Python (version 3.10.3) and R (version 4.5.1). Characterization of sncRNAs was conducted with the seqBuster/seqCluster^32,33^. Before starting, adaptors were trimmed with Cutadapt 3.5^34^, and reads from the same sample obtained from different runs (different FASTQ files) were collapsed. Before initiation quality control (QC), 23 samples were removed from the dataset due to sex discordance (n = 7), contamination from maternal DNA or sample manipulation (n = 7), identified through EPIC array DNA methylation data^35^, absence of DNA methylation data (n = 6), very low sncRNA sequencing depth (n = 2), and sampling from fetal membranes (n = 1). After these exclusions, the initial number of samples (including duplicates) was 457 (Appendix A: Fig. S1).

#### 2.3.1. Identification and quantification of miRNAs with seqBuster

SeqBuster was used for the identification and quantification of miRNAs. Briefly, reads were mapped to hairpins released from miRBase v22.1 (GRCh38)^36^ using miraligner, and sequences mapping ambiguously to several positions of the genome were filtered out. The average number of reads mapping to miRNAs was 0.83 M (min = 855, max = 1.65 M), representing 13.88% of the total number of reads (min = 0.02%, max = 24.85%). Samples with <0.3 M miRNA reads (n = 40) and technical duplicates (n = 27) were removed. For technical duplicates we kept the sample with the higher percentage of mapped reads (Pearson correlation of technical duplicates ranged from 0.91 to 0.99). After that, the 785 initial miRNAs identified were normalized to counts per million (CPMs) and those not showing at least 5 CPMs in at least 10% of the samples were excluded (n = 363). Then, miRNA expression was scaled with the trimmed mean of m values (TMM) method and log2 transformed with the edgeR tool^37^, ending up with a dataset of 390 unique samples and 422 log_2T_MM miRNAs. The transcriptome abundance 50 (TA50) metric, which represents the minimum percentage of features required to reach 50% of the total annotated miRNA reads, was calculated as an indicator of the diversity of expression within the placenta miRNAome.

The impact of technical variables (RIN, library preparation), biological variables (sex, gestational age, ethnicity and placental cell type proportions) and design variables (hospital of delivery and COVID-19 confinement) on miRNA levels was explored using Principal Component Analysis (PCA) and the variancePartition^38^ tool. To minimize technical bias, ComBat^39^ was applied to correct for the library preparation batch effects. Furthermore, we employed Surrogate Variable Analysis (SVA) ^40^ to account for latent technical factors, after residualizing the effect of primary biological or design variables. The first five surrogate variables (SVs), which collectively explained 50% of the variance, were included as technical covariates in the final models.

#### 2.3.2. Identification and quantification of sncRNAs with seqCluster

We used seqCluster to characterize and quantify sncRNAs beyond miRNAs. The tool handles sequence redundancy and multi-positional mapping by identifying sncRNA clusters, defined as groups of overlapping sequences that can be considered as single transcriptional units regardless of their annotation to the genome. This approach ensures that reads mapping to multiple genomic regions are not excluded, providing a more comprehensive view of the placental sncRNA landscape.

Briefly, sequences were aligned to the human reference genome GRCh38 using the STAR aligner^41^, allowing any number of multimap alignments. Then, seqCluster detected hotspots (sets of overlapping sequences according to their position in the genome), which were grouped in primary sncRNA clusters according to their genomic sequence. Next, a recursively heuristic algorithm based on reduction and cluster correction was applied to the ones sharing over 60% of the sequences. Finally, sncRNA clusters were annotated to different RNA species using miRbase^36^ v22.1 (GRCh38), refGene (RefSeq genes), wgRna (C/D and H/ACA Box snoRNAs and miRNAs), rmsk (Repeating elements created using RepeatMasker), tRNA from University of California Santa Cruz (UCSC) genome browser (GRCh38)^42^,and piRNAs from pidb 1.7.6 (GRCh38) ^43^.

Approximately 85% of reads were annotated as sncRNAs in the sncRNA dataset (min = 3.28 M, max = 6.83 M, mean: 4.89 M). Among the 457 initial samples, a total of 67 were removed: 38 with <3 M reads and <60% mapping to sncRNAs, 27 technical duplicates (similar to miRNAs), and two samples that could not be matched with the miRNA dataset. Among the 1,148 sncRNA clusters, 169 sncRNA were filtered out as they did not reach a minimum of 8 counts (representing 1.6 CPM cutoff) in at least 10% of the samples. Hence, the final dataset included 390 unique samples and 979 sncRNA clusters. We then normalized sncRNA levels using TMM method and applied a log2 transformation. As for miRNAs, we used ComBat to correct for the library preparation batch and additionally estimated SVs with the SVA method ^44,45^. We also selected 5 SVs as covariates, as explained up to 50% of the variance. Following the same approach used for the miRNAome, we calculated the TA50 of the sncRNAome.

Due to the underlying rationale of seqCluster, a single sncRNA cluster can map to multiple genomic loci and, consequently, be assigned to various sncRNA biotypes. To ensure consistent and interpretable annotation we established a biotype prioritization hierarchy. This hierarchy based on the a priori expected prevalence and potential functional relevance of sncRNA classes: miRNA, pre-miRNA, piRNA, tRF, snRNA, snoRNA, lncRNA, rRNA, protein-coding gene, pseudogene, and unknown (unannotated)^46^.

### 2.4. MiRNA nomenclature harmonization and cross-pipeline comparison

To enable direct comparisons between datasets, miRNAs were analyzed at the precursor level due to differences in annotation sources and naming conventions. The miRNA dataset was annotated using miRBase v22.1 (providing mature miRNA annotations, e.g., miR-103a-5p), while the sncRNA dataset used UCSC Genome Browser gene annotations (providing precursor gene nomenclature, e.g., *MIR103A*). Mature miRNA annotations from seqBuster were converted to precursor nomenclature by removing the mature-arm suffixes (-3p and -5p), while retaining family-specific lettered and numbered suffixes (e.g., miR-103a-1 to *MIR103A1*, where letters denote family members and numbers different genomic regions). miRNA expression concordance between the two pipelines (seqBuster at the mature level and seqCluster at the precursor level) was then assessed using Pearson correlation coefficients.

### 2.5. External comparison of the placental sncRNA profile

To compare the placental sncRNA profile identified in our study with previously published data, we used two studies: Gong *et al*. (2021), which characterized the placental RNA landscape using data from Pregnancy Outcome Prediction study (POPs), from which we specifically analyzed the miRNAs, snoRNAs and piRNAs, and Martínez *et al*. (2021), which specifically characterized placental piRNAs. We evaluated the overlap of detected sncRNAs between studies. Specifically, we assessed whether both all detected sncRNAs and the subset of top-expressed ones identified in our study were present in the published datasets, and vice versa.

### 2.6. Exposure assessment

#### 2.6.1. Prenatal air pollution exposure

In BiSC, maternal exposure to NO_2,_ PM_2.5,_ and BC was assessed using three different types of models: land use regression (LUR) models, dispersion models, and hybrid LUR-dispersion models. The details of these models have been described elsewhere^47^. Current study is based on the hybrid LUR-dispersion model, which showed better performance than the others and provides estimates of air pollution exposure at home and workplace.

Briefly, to develop the LUR models, we conducted four air pollution monitoring campaigns (2021-22) in 34 locations within the study area following the guidelines established by the European Study of Cohorts for Air Pollution Effects^48,49^. The dispersion models were developed using the ADMS-Urban software (Cambridge Environmental Research Consultants, Cambridge, UK), which is based on a Gaussian dispersion model with integrated photochemical reaction models, street canyon models, and a meteorological preprocessing model^50^. The hybrid models were developed using a random forest algorithm that combines the dispersion model outputs with the predictors from the LUR model together with both the routine air pollution monitoring and meteorological data. We estimated pollution-specific exposure levels for each participant for her entire pregnancy period as the average of exposure level at home and workplace, weighted by the time that the participant spent in each microenvironment. We applied a combination of a GPS-enabled smartphone app (ExpoApp - Smartphone app, Ateknea Solutions, Spain) and questionnaires to extract data on average time that each participant spent in each microenvironment.

#### 2.6.2. Maternal and partner tobacco smoking during pregnancy

Information on maternal smoking habits was self-reported at both the 12-week and 32-week visits. Based on that, mothers were classified as: non-smokers, smokers at early pregnancy (non-sustained smokers) and smokers during the whole pregnancy (sustained smokers). For the main analysis sustained and non-sustained smokers were grouped together (any maternal smoking) and the reference group were mothers who did not smoke during pregnancy (non-smokers).

Information on partner smoking habits was self-reported at the 20-week visit. Partners were classified as smokers if they reported active smoking during pregnancy, regardless of the number of cigarettes smoked or the frequency of smoking; otherwise were considered non-smokers.

### 2.7. Covariates

We collected data on potential confounders and covariates through follow-up visits, questionnaires, and medical records. The primary and sensitivity analyses included maternal age, maternal educational level, maternal body mass index (BMI), parity, hospital of delivery, neighborhood socioeconomic status (SES), offspring’s sex and ethnicity, gestational age at birth, season of conception, COVID-19 pandemic, and placental cell type proportions.

Maternal age was self-reported during the 12-week visit. Maternal BMI at 12 weeks was calculated as weight (kg) divided by height^2, both measurements obtained during the 12-week visit. Parity (nulliparous/multiparous) was obtained from medical records. Hospital of delivery was classified into 3 categories: HSjD, HSP and other (including Hospital Clínic-Maternitat, home births and other hospitals).

Maternal education level (university/non-university) was self-reported and included in our analysis as a surrogate for individual-level SES. As a proxy of neighborhood SES, we used the annual average household income (euros) at census tract level obtained from the 2020 Standard of Living and Living Conditions survey by the Spanish National Institute of Statistics^51^.

Offspring sex (female/male) was obtained from medical records and validated through molecular analysis. Child ethnicity (European/other) was obtained from self-reported maternal and paternal ethnicity; children were classified as "European" if both parents were of European descent, and "other" otherwise. Birth weight was obtained at delivery from medical records and gestational age at birth was calculated from the date of the last menstrual period using estimates based on the first ultrasound examination (around 12th week of gestation)^52^. Preterm was defined as a gestational age at birth <37 completed weeks. Information on obstetric complications (pre-eclampsia/eclampsia, gestational hypertension, placental abruption, placenta previa, oligohydramnios, polyhydramnios and chorioamnionitis) was obtained from medical records. The proportions of placental cell type populations, including trophoblasts, syncytiotrophoblast, nucleated red blood cells (nRBCs), Hofbauer cells, endothelial cells, and stromal cells, were estimated from DNA methylation data measured in the same placental biopsy using the placental reference panel described in the planet R package^53^ .

Season of conception (autumn/spring/summer/winter) was obtained by subtracting the gestational age from the date of birth. As the pregnancy period of some of the women of the BiSC cohort overlapped with the COVID-19 pandemic which affected lifestyle and the protocols of sample collection in hospitals, we created a variable indicating whether the pregnancy took place before, during, or after the confinement, and the confinement was defined between March 14th 2020 (WHO declaration of Global Pandemic) and May 9th 2021 (end of the alarm state in Spain).

### 2.8. Statistical analyses

#### 2.8.1. Descriptive analyses

Descriptive statistics for covariates among the selected participants (n = 390) and the complete-case subset on exposures, covariates, and sncRNA profiles (n = 359) were generated using the tableone^54^ package in R. Comparisons between the selected participants and the remaining cohort (n = 690 mothers; n = 642 children) were conducted using the same package. Statistical significance was evaluated using Mann-Whitney U test for continuous variables and chi-square tests for categorical variables. The standardized mean difference (SMD) was also calculated to quantify the magnitude of differences between groups.

Correlation across exposures was examined using the Pearson’s correlation for continuous variables (BC, NO_2,_ and PM_2.5)_ and the point-biserial correlation for combinations of categorical (smoking status) and continuous variables.

#### 2.8.2. Differential sncRNA expression analyses

The differential expression analyses of placental sncRNAs in relation to pregnancy exposures were conducted with the limma^55^ R package. A linear regression model was fitted for each feature (miRNA or sncRNA cluster) and each exposure variable (BC, NO_2 o_r PM_2.5 e_xposure levels and MSDP), adjusting for the child’s sex, ethnicity, gestational age at birth, parity, maternal education, maternal BMI at recruitment, maternal age, COVID-19 confinement, hospital of delivery, season of conception, neighborhood SES, placental cell type proportions, and 5 SVs.

Multiple-testing correction was addressed with the effective number of independent tests (ENT) method^56^. The ENT was calculated based on the total number of features (1,401, including both miRNAs and sncRNA clusters) and the four exposures (BC, NO_2,_ PM_2.5,_ and MSDP). ENT was 272.37 for features and 2.50 for exposures, giving an adjusted p-value < 7.34 × 10^-5^ [0.05 / (272.37 x 2.50)]. As suggestive statistical significance we used a p-value < 1.836 × 10^-4^ calculated considering the features and not the exposures (0.05 / 272.37).

The effect sizes are reported as the log2 fold change (log2FC) in miRNA or sncRNA levels. For air pollution, this represents the change per interquartile range (IQR) increase in exposure; for MSDP, it represents the difference between smokers and non-smokers. The results were graphed with Volcano plots using ggplot2^57^ R package.

Five sensitivity analyses were conducted: (i) restricted to participants of European ethnicity, the largest ethnic subgroup in the cohort; (ii) excluding participants with obstetric complications (preterm birth and pregnancy complications related to placental dysfunction, including preeclampsia/eclampsia, gestational hypertension, placental abruption, placenta previa, oligohydramnios, polyhydramnios, and chorioamnionitis); (iii) without adjusting for gestational age; (iv) without adjusting for COVID-19 confinement; and (v) additionally adjusting for partner’s smoking during pregnancy (only for the MSDP models).

### 2.9. Downstream in silico analyses

#### 2.9.1. Biological characterization of exposure-associated miRNAs

We conducted downstream analyses of the top five ranked miRNAs associated with each exposure identified by seqBuster and/or sncRNA clusters for which the prioritized RNA type was a miRNA. To characterize the biological relevance of these miRNAs, we investigated their predicted and experimentally validated target genes. Predicted miRNA-gene interactions were obtained from miRDB-v6.0 (https://mirdb.org/), from which the five highest-ranked target genes with prediction scores >90 were selected. Experimentally validated miRNA-gene interactions were retrieved from TarBase-v9.0 (https://dianalab.e-ce.uth.gr/tarbasev9), selecting the five target genes supported by the strongest experimental evidence.

To determine whether target genes exhibited placental-enriched expression, we interrogated the POPs placental transcriptome database reported by Gong *et al*. (2021) (https://www.obgyn.cam.ac.uk/placentome/). Genes were considered placental-enriched if they were expressed at ≥0.1 transcripts per million (TPM) in placenta and exhibited expression levels at least 10-fold higher than the average across 49 Genotype-Tissue Expression (GTEx) tissues. Additionally, protein expression of target genes was retrieved from the Human Protein Atlas (HPA; https://www.proteinatlas.org/). The dataset was filtered to retain only protein expression annotations with “Approved”, “Enhanced”, or “Supported” reliability scores. For each target gene, protein expression levels were assessed across normal tissues and specifically within placenta. Expression annotations were categorized according to HPA protein abundance levels (high, medium, or low) and, when available, assigned to specific placental cell types. For genes lacking available protein expression annotations across normal tissues or within placental cell types after filtering, expression was classified as “No protein expression reported” or “No placental protein expression reported”, respectively.

#### 2.9.2. Characterization of the *DLK1-DIO3* locus

Characterization of the *DLK1*-*DIO3* locus was performed at multiple levels. First, we assessed annotation coverage by comparing the miRNA, piRNA, and snoRNA annotations detected in our dataset with those previously reported for the *DLK1*-*DIO3* locus. Second, we evaluated genomic overlap between snoRNA and piRNA annotations to identify regions with dual annotation using the *GenomicRanges* ^58^ package in R. Genomic coordinates were obtained from piRNAdb v1.7.6 and the UCSC wgRna annotation track for snoRNAs.

To study the sdRNAs, sequences belonging to each snoRNA cluster were collapsed into representative sequence families based on identical genomic coordinates (chromosome, start, and end positions). For each family, average CPM was calculated and the most abundant sequence was retained as the representative sequence. Low-abundance sequence families (average CPM ≤ 0.01) were excluded from further analyses. The genomic coordinates of the C, D, C′, and D′ box motifs were obtained from snoDB (https://bioinfo-scottgroup.med.usherbrooke.ca/snoDB/) and used to annotate the position of the representative fragments within each snoRNA. The genomic distribution of all representative sequence families across each snoRNA locus was graphed together with the annotated box motifs. The representative fragment accounting for the highest proportion of total cluster counts was selected for downstream structural analyses.

Two-dimensional secondary structures of the full-length snoRNAs were predicted using the RNAfold tool from the ViennaRNA Web Services (http://rna.tbi.univie.ac.at/). For each snoRNA, the minimum free energy structure was selected for downstream analyses and visualized using Forna, also available through the ViennaRNA Web Services. Representative fragments were highlighted within the predicted secondary structures according to their positions within the snoRNA sequence to assess whether the observed molecules were consistent with specific snoRNA-derived fragments rather than random degradation products. The mature snoRNA sequence was obtained from snoDB.

## 3. Results

### 3.1. Study population

The study population consisted of 390 mother-child pairs from the BiSC cohort with available sncRNA profiles. Among this, 359 had complete data on exposures, covariates, and sncRNA profiles and were included in the differential expression analysis (Appendix A: Table S1). The main characteristics of the 390 mother-child pairs included in the study are presented in Table 1. The study population was broadly comparable to the remainder of the cohort (Appendix A: Table S2). Small statistically significant differences were observed, mainly reflecting recruitment patterns during the COVID-19 pandemic, when a greater proportion of placentas were collected at HSjD and ambient PM_2.5 c_oncentrations were lower during the confinement period.

**Table 1.**
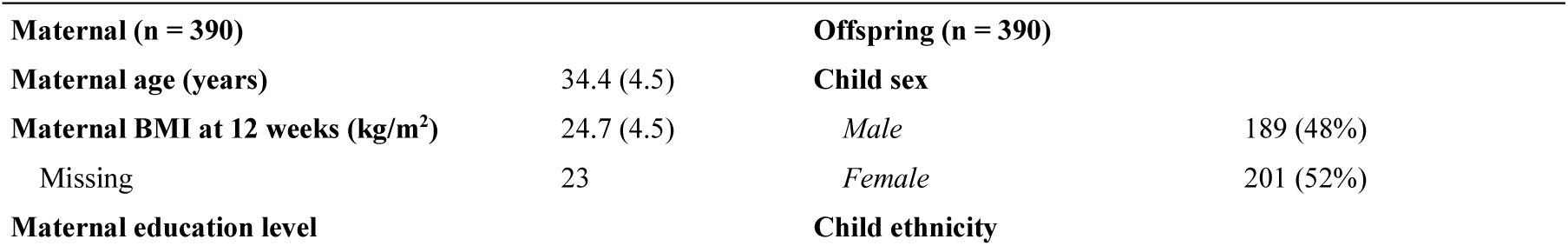
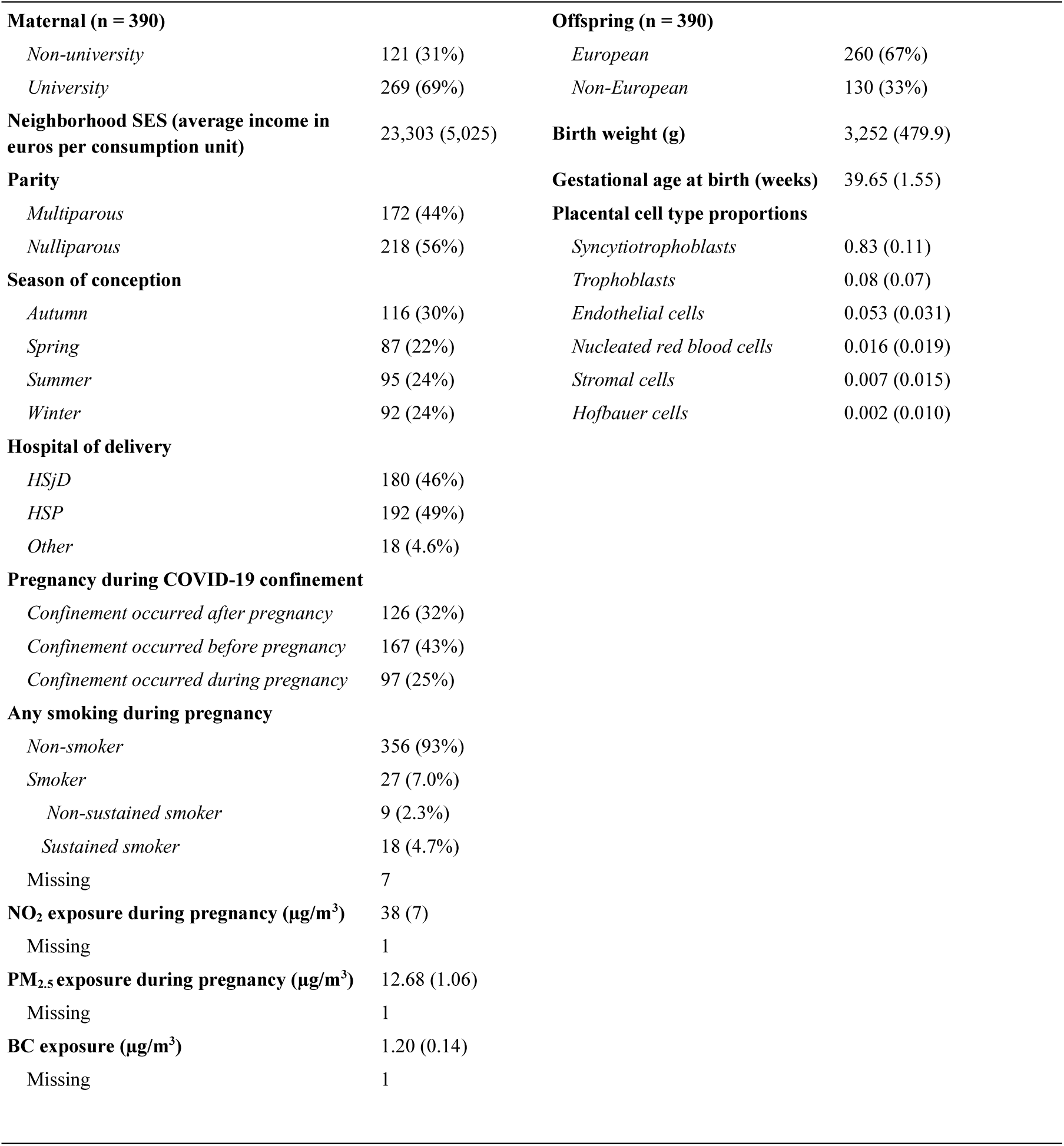
Descriptive characteristics of the mother-child pairs included in the study (n=390). Continuous variables are presented as mean ± standard deviation (SD), and categorical variables as number and percentage. *BC: black carbon; BMI: body mass index; HjD: Hospital Sant Joan de Déu; HSP: Hospital de Sant Pau; NO_2_: nitrogen dioxide; PM_2.5_: particulate matter with diameter ≤ 2.5 μm; SES: socioeconomic status*

The mean age of the mothers at delivery was 34.4 years (SD = 4.5), over half of them were nulliparous (56%), and 69% hold a university degree or higher. COVID-19 confinement occurred during pregnancy for 25% of the women. Forty-eight % the children were females, 67% were of European ethnicity, the average birth weight was 3,252 g (standard deviation, SD = 479.9), and the average gestational age was 39.65 weeks (SD = 1.55). Twenty-seven mothers (7%) smoked at some point during pregnancy, with 9 classified as early smokers (2.3%) and 18 as sustained smokers (4.7%). Regarding air pollution exposures, mean levels were 38 µg/m^3^ (SD = 7) for NO_2,_ 12.6 µg/m^3^ (SD = 1) for PM_2.5 a_nd 1.2 µg/m^3^ (SD = 0.14) for BC. Strong correlations were observed among the air pollutants (Pearson’s correlations 0.60 - 0.82), while MSDP showed no correlation with any of them (Appendix B: Fig. S2).

### 3.2. Placenta sncRNA profiles

Reference-based deconvolution of placental DNA methylation data yielded cell type proportions similar to those reported in previous population-based studies taking the biopsy from chorionic villi^59^. Syncytiotrophoblasts represented the predominant cell type (mean = 0.83, SD = 0.11), followed by trophoblasts, endothelial cells, nRBCs, stromal and Hofbauer cells.

Sequencing data was pre-processed with the seqBuster/seqCluster pipeline, producing a dataset of 422 miRNAs and a dataset of 979 sncRNA clusters (see Methods, section 2.3.1 and section 3.2.1. . One-hundred and ninety-two sncRNA clusters (19.6%) were annotated to a single biotype, 314 to two biotypes (32.0%), 339 to three biotypes (34.6%), and 134 to more than three biotypes (13.6%) (Fig. 1A). After applying the prioritization criteria for describing sncRNA annotation (see Methods, section 2.3.2), 33.2% of the sncRNA clusters were annotated to miRNAs, followed by 26.4% to piRNAs, 12.2% to snoRNAs and 7.8% to tRFs (Fig. 1B). A small proportion of clusters (4.2%) remained unannotated, while the remaining 1.6% were annotated to pseudogenes, rRNAs, and snRNAs. Finally, 10.0% of the clusters overlapped genomic regions annotated as protein-coding genes and 4.2% as long non-coding RNAs (lncRNAs).

**Fig. 1.**
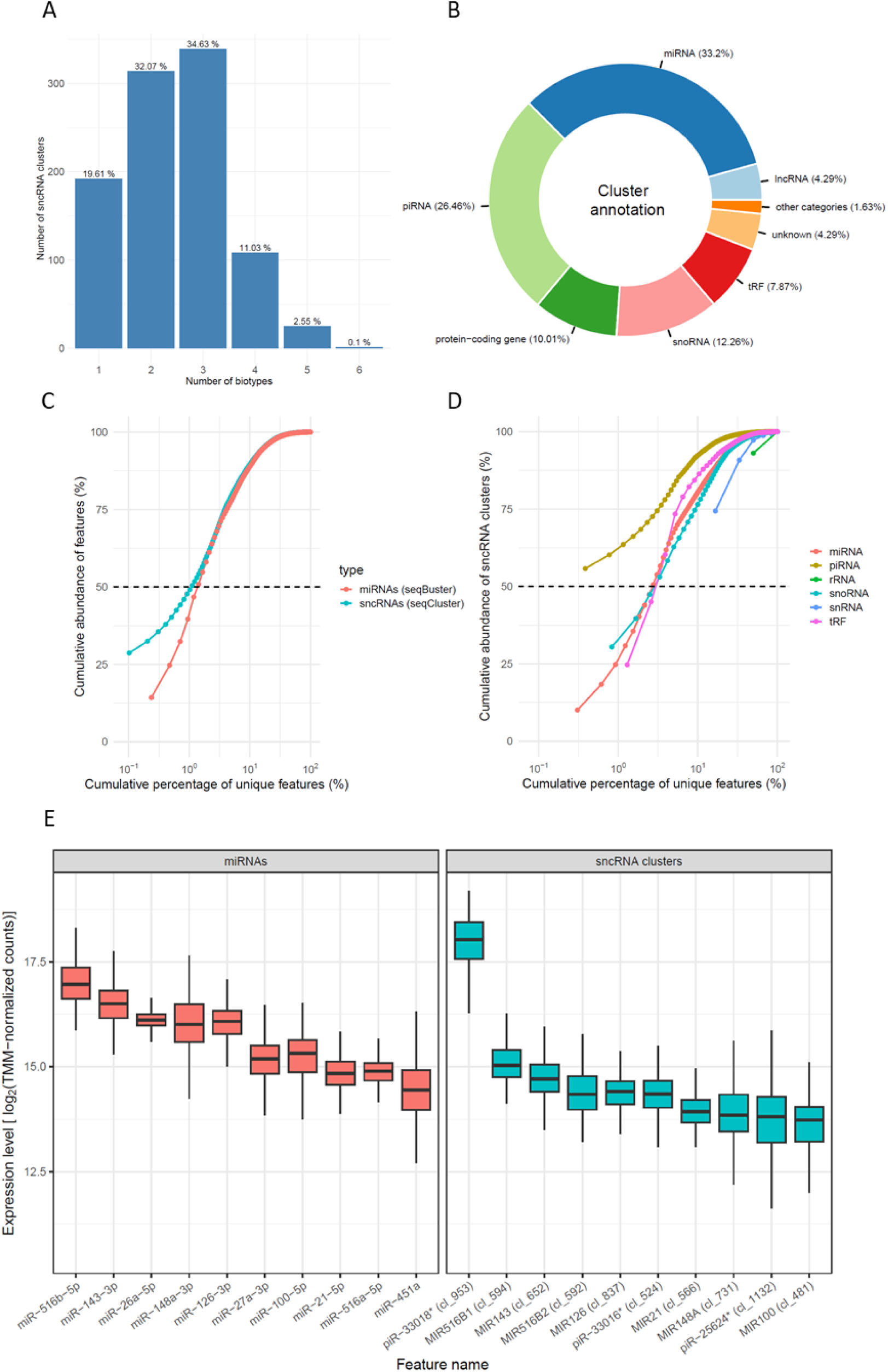
Placental miRNA and sncRNA cluster profiling. **A.** Distribution of the number of RNA types annotated per sncRNA cluster. The x-axis shows the number of distinct RNA types assigned to each sncRNA cluster, while the y-axis shows the number of sncRNA clusters. **B**. Pie chart summarizing distribution of sncRNA clusters according to the prioritization criteria: *miRNA, pre-miRNA, piRNA, tRF, snRNA, snoRNA, lncRNA, rRNA, protein-coding gene, pseudogene, and unknown (unannotated)*. “Other categories” include: pseudogene (0.82%), snRNA (0.61%), and rRNA (0.20%). **C-D.** Cumulative abundance (TA50) plots for miRNAs (seqBuster; red) and sncRNA clusters (seqCluster; blue). The x-axis shows the cumulative percentage of unique features within the total dataset (**C**) or within each specific RNA type (**D**). The y-axis represents the cumulative percentage of total annotated reads. The horizontal dashed line marks the 50% abundance threshold (TA50). **E**. Expression levels of top 10 most abundant miRNAs (red) and sncRNA clusters (blue). Boxplots represent expression levels as log2 TMM-normalized counts across the cohort samples. For sncRNA clusters, the prioritized RNA type is shown followed by the sncRNA cluster ID in brackets. The asterisk (*) indicates that the sncRNA cluster contains multiple annotations of the same RNA type; in these cases, only the first feature is displayed.

After harmonizing miRNA annotations to the precursor level, a total of 362 miRNA genes were identified across the seqBuster and seqCluster outputs. Of these, 271 (74.8%) were detected by both pipelines. Expression estimates for shared miRNAs showed a strong correlation between pipelines, with an average Pearson correlation coefficient of r = 0.74 (min = 0.71, max = 0.77). When the analysis was restricted to the 51 seqCluster clusters exclusively annotated as miRNAs, concordance increased further, with an average correlation of r = 0.79 (min = 0.73, max = 0.84).

To examine expression dominance, we calculated the cumulative contribution of miRNAs and sncRNA clusters to the corresponding overall mapped RNA pool (Fig. 1C). The TA50, representing the minimum percentage of unique features required to account for 50% of the total expression, was comparable across both datasets. Specifically, only five unique miRNAs (representing 1% of the total detected miRNAs) were sufficient to reach half of the total miRNA reads. Similarly, for the sncRNA cluster dataset, 10 clusters (representing 1% of all identified clusters) accounted for 50% of the total abundance. Additionally, within the sncRNA dataset, the number of features required to reach TA50 varied across RNA types: snoRNAs (n = 3, 2.5%), tRFs (n = 2, 2.5%), and miRNAs (n = 8, 2.4%). For piRNA, rRNA, and snRNA, the TA50 value was effectively zero because a single feature accounted for >50% of the total RNA type abundance (Fig. 1D; Appendix A: Table S3).

Analysis of the most abundant features (Fig. 1E) revealed substantial overlap between the miRNA and sncRNA datasets. Seven of the 10 most abundant mature miRNAs were also among the 10 most abundant sncRNA clusters, which were annotated to their corresponding precursor miRNA loci. Notably, miR-516b-5p, the most abundant miRNA detected, and miR-516a-5p, which also ranked among the top 10 miRNAs, belong to the placenta-enriched C19MC cluster. Other miRNA members of the placental predominant clusters were also detected in our samples: 78.6% (33/42) of C14MC miRNAs, 80.4% (37/46) of C19MC miRNAs, and 66.7% (2/3) of miRNAs from the miR-371-373 cluster.

Apart from miRNAs, piRNAs represented the second most abundant RNA class in the sncRNA dataset. Specifically, the most abundant cluster, cl_953, was annotated to 20 distinct piRNAs, while cl_524 and cl_1132 were annotated to three and 12 piRNAs, respectively.

To assess the consistency of our findings, we evaluated whether the RNA types identified in our study had also been reported in previous placental sncRNA studies. Specifically, we compared our results with the placental sncRNA transcriptome described by Gong *et al*. (2021). We observed substantial overlap across miRNAs as 97.4% (411/422) of mature miRNAs (seqBuster) and 98.6% (357/362) of precursor miRNAs (seqCluster) identified in our study were also reported by Gong *et al*. Likewise, 92.7% (190/205) of snoRNAs and 60.7% (297/489) of piRNAs were shared between the two studies. Moreover, eight of our 10 most abundant mature miRNAs were also among most abundant placental miRNAs reported by Gong *et al*., including miR-516b-5p, miR-143-3p, miR-26a-5p, miR-148a-3p, miR-126-3p, miR-27a-3p, miR-100-5p, and miR-21-5p. Similarly, SNORD2, one of the most abundant snoRNAs in their study, was also detected within our highly expressed cluster cl_524.

We also compared our piRNA results with those reported by Martínez *et al*. (2021). Only 8.0% (24/297) of their consistently expressed piRNAs were identified in our study. This subset included piR-28398, which was detected within our most abundant piRNA-containing cluster (cl_953). However, none of the placenta-specific piRNAs reported by Martínez *et al*. were detected in our study.

### 3.3. Differential expression analysis of air pollution exposure and tobacco smoking during pregnancy

To investigate the associations of prenatal air pollution exposure (BC, NO_2,_ PM_2.5)_ and MSDP with placental levels of miRNAs and sncRNA clusters, we fitted linear regression models adjusted for potential confounders. After multiple-testing correction (p-value < 7.34 x 10^-5^), we found that MSDP (comparing *any smoking* to *non-smokers*) was associated with the downregulation of miR-421 (Fig. 2A; Table 2) and the upregulation of two sncRNA clusters (cl_201 and cl_207) located at the imprinted *DLK1-DIO3* locus (Fig. 2B; Table 3). At the suggestive significance threshold (p-value < 1.84 x 10^-4^), two additional upregulated sncRNA clusters (cl_193 and cl_214) were identified, both at the same *DLK1-DIO3* locus. In addition, cl_196, which was marginally significant (p-value = 2.29 x 10^-4^), was also located within this locus.

**Fig. 2.**
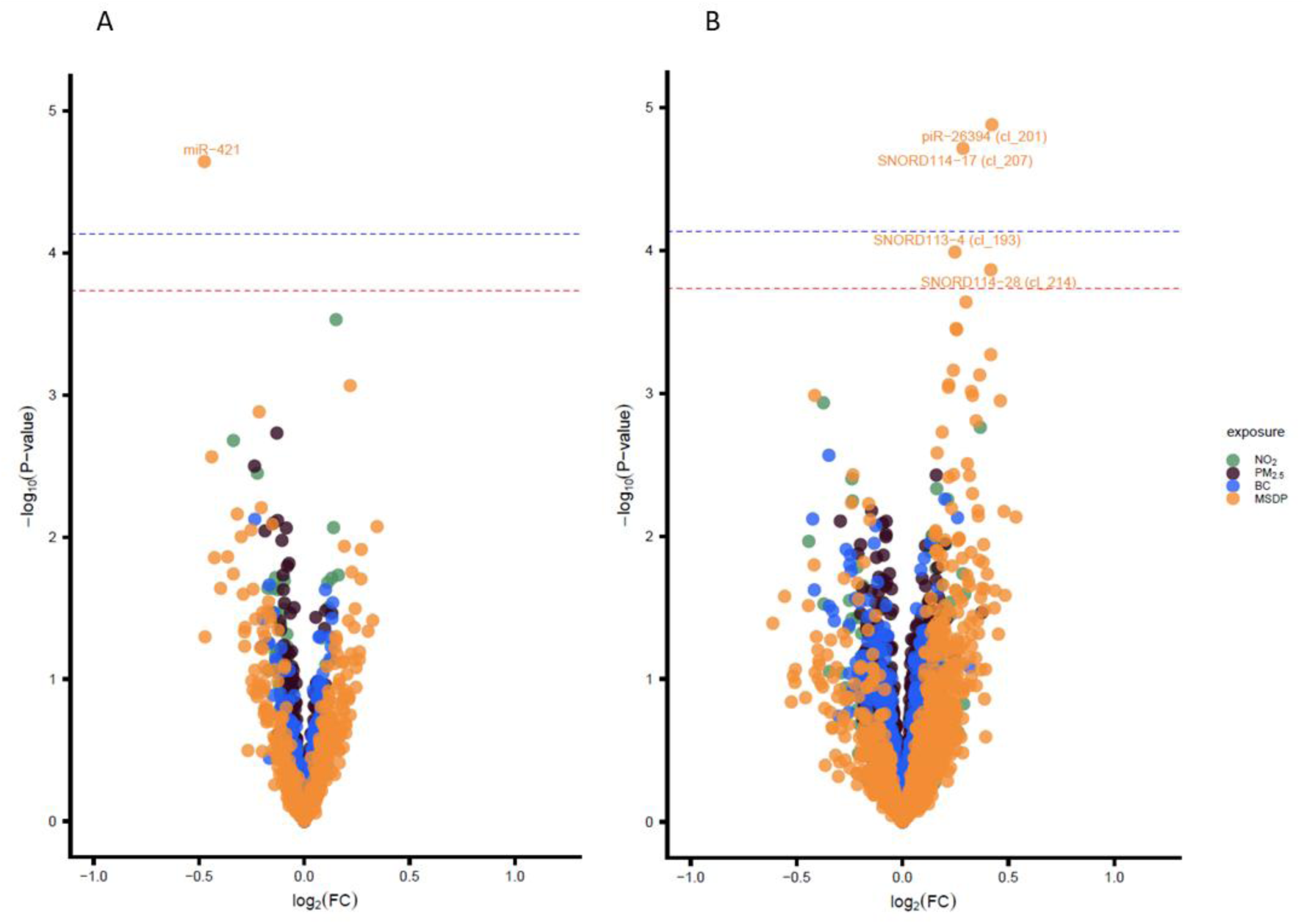
**A-B.** Volcano plots illustrating the associations between each exposure (BC, NO_2_, PM_2.5_, and MSDP) and placental levels of miRNAs (**A**) and sncRNA clusters (**B**). The x-axis represents association coefficients (log_2_FC per IQR increase for air pollutants, and for “any smoking” vs “non-smoking” for MSDP), and the y-axis shows the statistical significance (-log10 p-value). Each point corresponds to a feature (miRNA or sncRNA cluster), with colors indicating the exposure: NO_2_ (green), PM_2.5_ (black), BC (blue), and MSDP (orange). Horizontal lines indicate significance thresholds (ENT: significant: p-value < 7.34 × 10^-5^, blue line; suggestive: p-value <1.84 × 10^-4^, red line). *BC: black carbon; NO_2_: nitrogen dioxide; MSDP: maternal smoking during pregnancy; PM_2.5_: particulate matter with diameter ≤ 2.5 μm*.

**Table 2.**
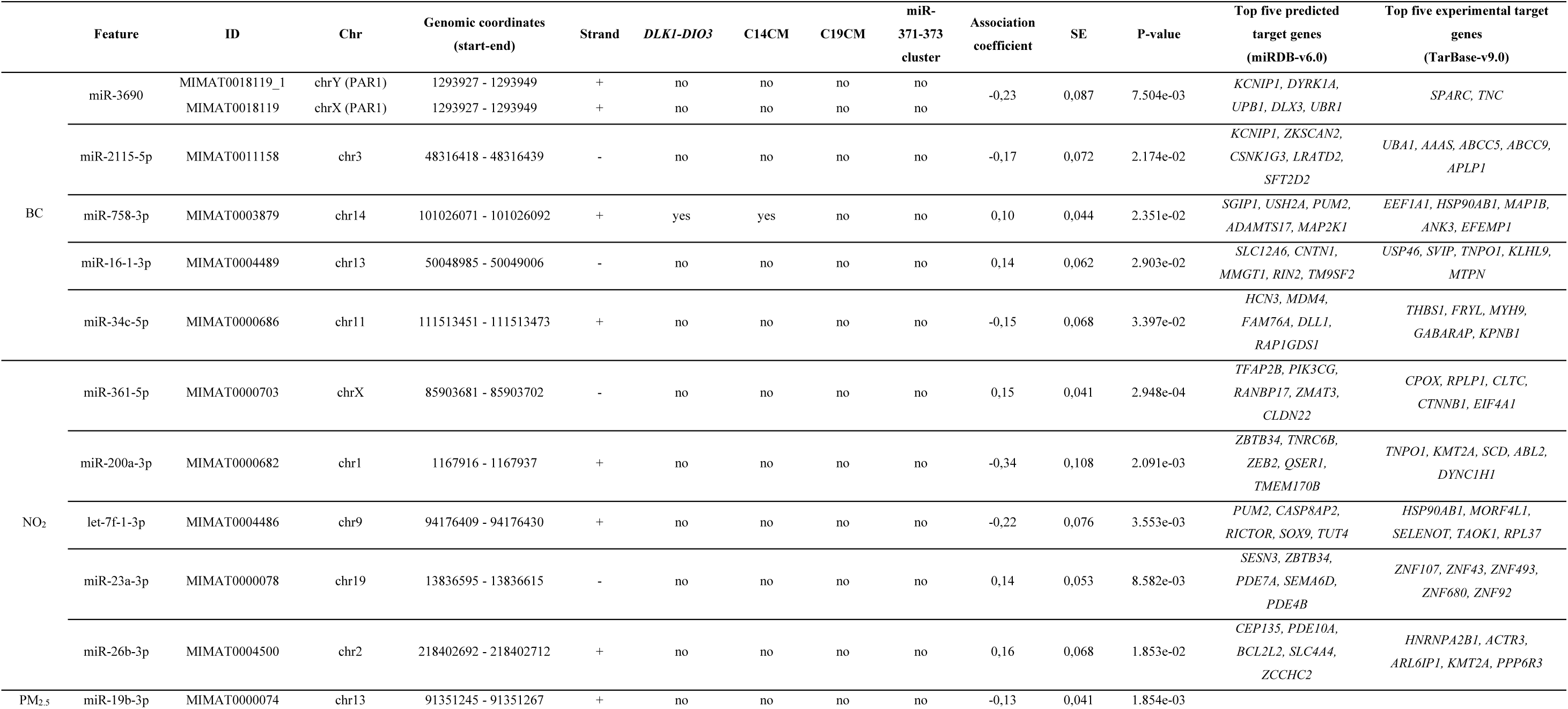
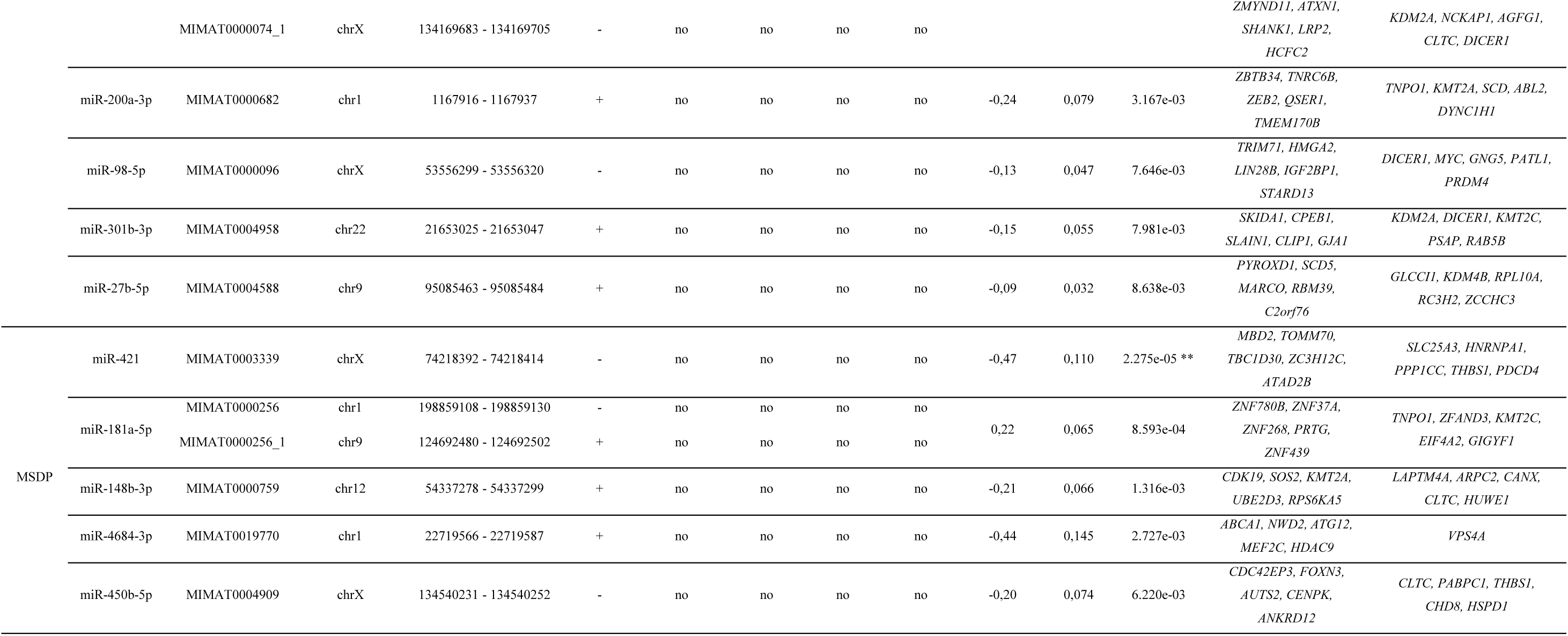
Summary of the five most statistically significant miRNAs for each exposure (BC, NO_2_, PM_2.5_, and MSDP) based on the main association models (n = 359). The table reports the genomic position of each miRNA based on the GRCh38 assembly, including chromosome (Chr), genomic coordinates (start-end), strand orientation (+: sense; -: antisense) and whether the miRNA belongs to the placental miRNA clusters C14MC, C19MC, and miR-371-373. It also includes the main model association coefficient, standard error (SE), and p-value, together with the top five predicted and experimentally validated target genes according to miRDB-v6.0 and TarBase-v9.0, respectively. Significance according to ENT-adjusted p-value thresholds is indicated as follows: * p-value < 1.84 × 10^-4^ (adjusted by total number of features across both RNA-seq datasets); ** p-value < 7.34 × 10^-5^ (adjusted by total number of features across both RNA-seq datasets and total number of exposures). *BC: black carbon; Chr: chromosome; C14CM: chromosome 14 miRNA cluster; C19CM: chromosome 19 miRNA cluster; MSDP: maternal smoking during pregnancy, NO_2_: nitrogen dioxide; PAR1: pseudoautosomal region 1; PM_2.5_: particulate matter with diameter ≤ 2.5 μm;*

**Table 3.**
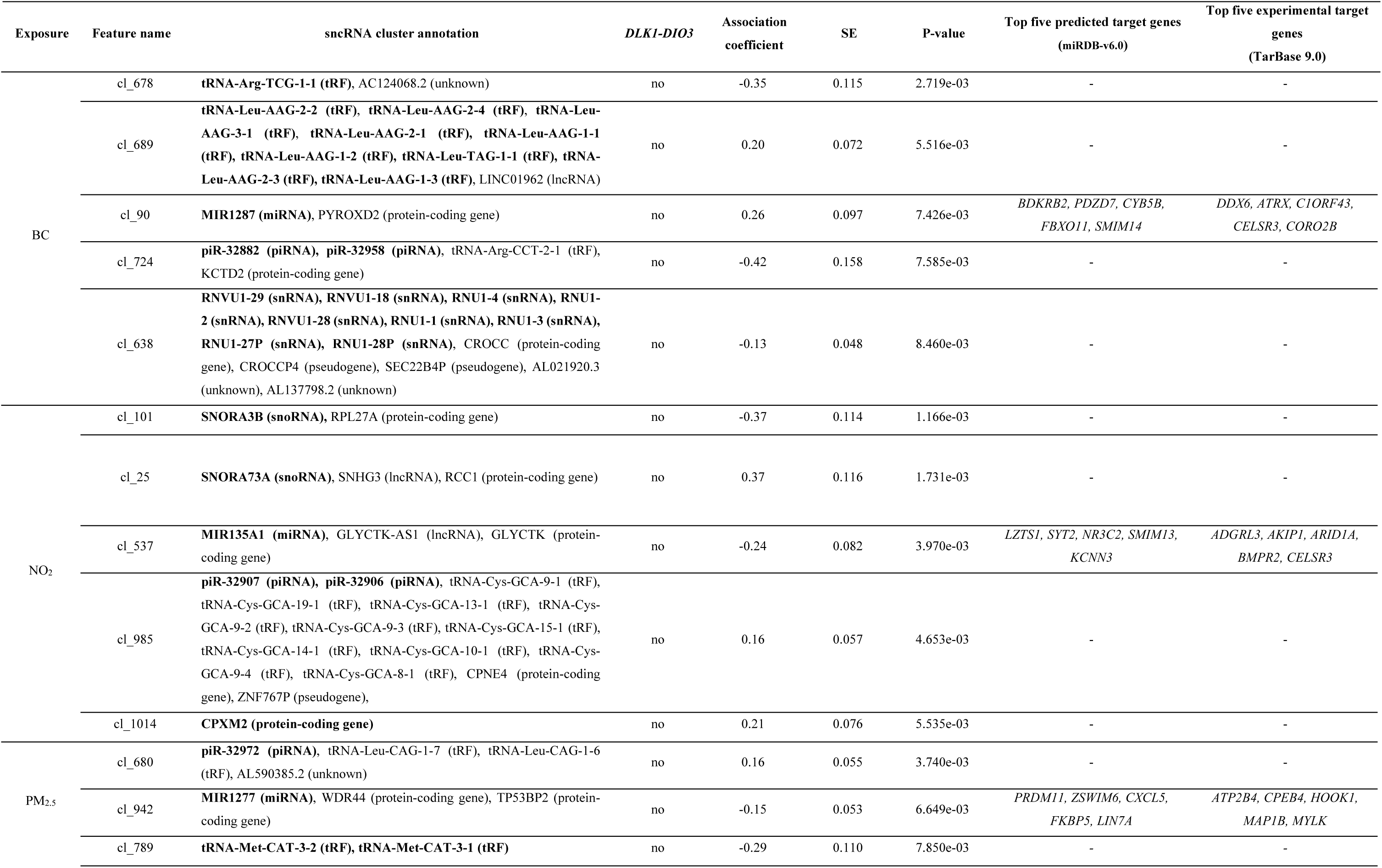
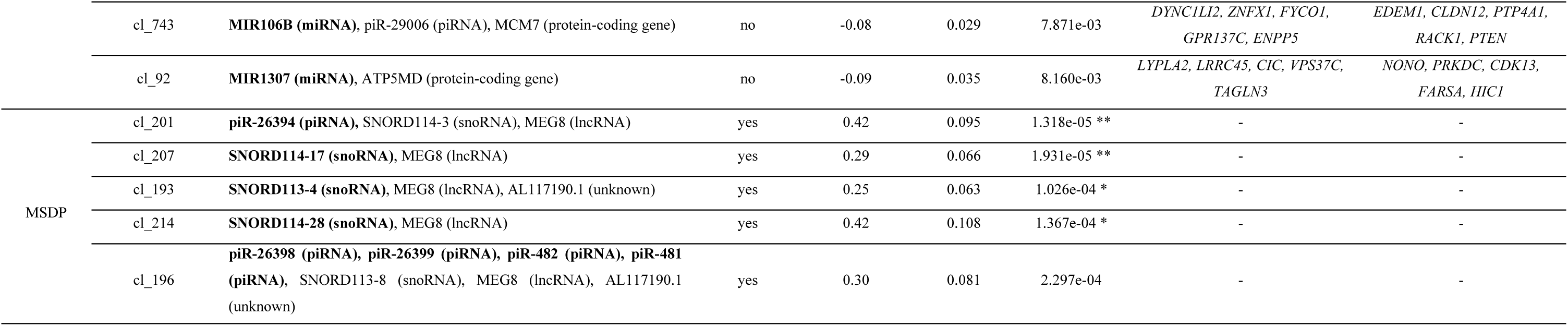
Summary of the five most statistically significant sncRNA clusters for each exposure (BC, NO_2_, PM_2.5_, and MSDP) based on the main association models (n = 359). The table includes the sncRNA cluster annotation (prioritized RNA type marked in bold), whether the sncRNA cluster belongs to the *DLK1-DIO3* locus, the association coefficient, standard error (SE), and p-value, along with the top five predicted and experimentally validated target genes according to miRDB-v6.0 and TarBase-v9.0, respectively, for sncRNA clusters whose prioritized RNA type is a miRNA. Significance according to ENT-adjusted p-value thresholds is indicated as follows: * p-value < 1.84 × 10^-4^ (adjusted by total number of features across both RNA-seq datasets); ** p-value < 7.34 × 10^-5^ (adjusted by total number of features across both RNA-seq datasets and total number of exposures). *BC: black carbon; MSDP: maternal smoking during pregnancy, NO_2_: nitrogen dioxide; PM_2.5_: particulate matter with diameter ≤ 2.5 μm*

We further examined the expression levels of these five features in relation to MSDP duration (non-smokers, non-sustained smokers, sustained smokers). For some features (cl_201, cl_207, and cl_193), longer duration was associated with greater changes in expression (Appendix B: Fig. S3). Because miR-421 is located within the pseudoautosomal region 1 (PAR1) of chromosome X, we performed sex-stratified analyses; however, no significant differences between female and male placentas were observed (Appendix B: Fig. S4).

No miRNAs or sncRNA clusters were significantly associated with any of the air pollution exposures. The top five miRNAs and sncRNA clusters for each exposure are shown in Table 2 and Table 3, respectively; full results are provided in Appendix A: Table S4 and Table S5.

For both miRNAs and sncRNA clusters with a p-value < 0.05 in the main model, we conducted four sensitivity analyses (see Methods, section 2.8.2). Association coefficients for MSDP and air pollution exposures remained highly consistent across sensitivity analyses in both the miRNA and sncRNA datasets (Appendix B: Fig. S5 and Fig. S6, respectively). For the five smoking-associated features, we calculated the percentage change in the association coefficient between the main and each sensitivity model (Appendix A: Table S6 and Table S7). The model restricted to pregnancies without complications and without adverse birth outcomes (n = 312) showed the greatest increase in the coefficients, with percentage changes ranging from 17.7% to 26.9%, while the model additionally adjusted for partner’s smoking showed the largest decrease in the coefficients, with percentage changes ranging from -29.8% to -48.1%, likely due to largely reduced sample (n = 218). However, partner’s smoking was not significantly associated with any of the sncRNA clusters.

### 3.4. Identification of target genes for exposure-responsive miRNAs

We investigated the predicted and experimentally validated target genes of the five most strongly associated miRNAs and miRNA-prioritized sncRNA clusters across all exposures using miRDB-v6.0 and TarBase-v9.0, respectively (Table 2 and Table 3). Sixty percent of target genes identified for the exposure-responsive miRNAs and 42% of those identified in the sncRNA dataset were expressed at medium to high protein levels in placental cell types cells according to the HPA (Appendix A: Table S8 and Table S9). Among all identified, only *LIN28B*, a predicted target of miR-98-5p (PM_2.5 e_xposure, p-value = 7.64 x 10^-3^), was classified as placenta-enriched (TPM ≥ 0.1 and expression ≥10-fold higher than the average across 49 GTEx tissues) according to the POPs^26^ placental transcriptome database.

### 3.5. Smoking associated sncRNA clusters located within the *DLK1-DIO3* imprinted locus

Given that top smoking-associated sncRNA clusters (cl_201, cl_207, cl_193, cl_214, and cl_196) mapped to the *DLK1-DIO3* locus, we further characterized this imprinted genomic region (Fig. 3). The *DLK1-DIO3* locus is located on chromosome 14q32 and contains several paternally expressed protein-coding genes (*DLK1*, *RTL1* and *DIO3*) and a large, maternally expressed polycistronic transcription unit. This single transcript encompasses lncRNA genes such as *MEG3*, *RTL1as*, and *MEG8,* which are processed to release multiple nested snoRNAs and miRNAs.

**Fig. 3.**
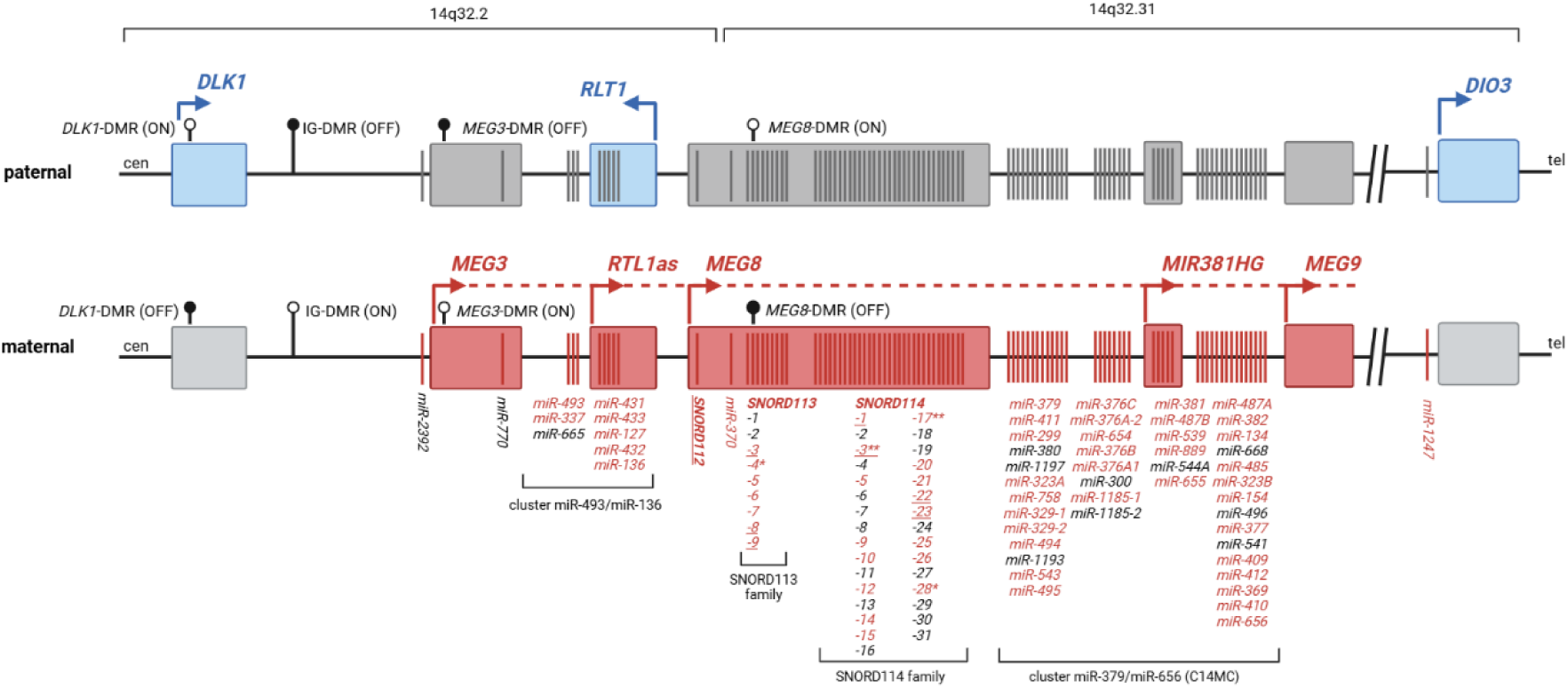
The *DLK1*-*DIO3* imprinted region contains the paternally expressed protein-coding genes *DLK1*, *RTL1*, and *DIO3* (blue), together with the maternally expressed long non-coding RNA genes *MEG3*, *MEG8*, *MEG9*, and *RTL1as* (red). The locus also harbors large maternally expressed clusters of sncRNAs, including miRNAs and snoRNAs. *MEG8* hosts tandemly repeated C/D-box snoRNA clusters comprising SNORD112 (1 copy), SNORD113 (9 copies), and SNORD114 (31 copies). Two major miRNA clusters, located at 14q32.2 (miR-493/miR-136 cluster) and 14q32.31 (C14MC), are also maternally expressed. Differentially methylated regions (DMRs) are indicated, including the *DLK1*-DMR, the intergenic IG-DMR, the *MEG3* promoter DMR, and the *MEG8*-DMR. DNA methylation is represented by lollipop symbols, where filled circles indicate methylated CpGs and open circles indicate unmethylated CpGs. The IG-DMR and *MEG3*-DMR are methylated on the paternal allele, whereas the *DLK1*-DMR and *MEG8*-DMR are methylated on the maternal allele. The dashed line represents the maternally expressed polycistronic transcript initiated from the *MEG3* promoter, which gives rise to multiple non-coding RNAs, including snoRNAs and miRNAs. Transcriptionally active genes are shown as colored elements, whereas inactive genes are represented in gray. Features detected in the present study are indicated in red text, and underlined snoRNAs indicate snoRNA loci that overlap annotated piRNAs. Not drawn to scale. Significance according to ENT-adjusted p-value thresholds is indicated as follows: * p-value < 1.84 × 10-4 (adjusted by total number of features across both RNA-seq datasets); ** p-value < 7.34 × 10-5 (adjusted by total number of features across both RNA-seq datasets and total number of exposures). Created in BioRender. *C14MC: chromosome 14 miRNA cluster; DMR: differentially methylated region* Adapted from: D’Angelo et al. (2026), and Baulina et al. (2021).

In our BiSC samples, we detected 77.8% (42/54) of the miRNAs annotated within this locus, including 87.5% (7/8) of miRNAs from the miR-493/miR-136 cluster, 78.6% (33/42) from the C14MC cluster, and two others not assigned to any specific cluster (miR-370 and miR-1247). We also detected several snoRNA from the C/D box class: *SNORD112* (1/1), 77.8% (7/9) of *SNORD113* family members, and 51.6% (16/31) of *SNORD114* family members. Furthermore, the analysis of snoRNA and piRNA genomic coordinates revealed that several snoRNAs within this locus overlap with piRNAs, specifically *SNORD112*, *SNORD113* (members 3, 8, and 9), and *SNORD114* (members 1, 3, 22, and 23) (Appendix A: Table S10).

All five smoking-associated sncRNA clusters mapped within the *MEG8* lncRNA gene and were upregulated in response to MSDP. Two clusters were annotated to both snoRNAs and piRNAs (cl_201: SNORD114-3 and piR-26394; cl_196: SNORD113-8 and piR-26398, piR-26399, piR-481, and piR-482), whereas the remaining three were annotated exclusively to snoRNAs (cl_207: SNORD114-17; cl_193: SNORD113-4; and cl_214: SNORD114-28).

Since some snoRNAs undergo enzymatic cleavage to yield smaller functional fragments (sdRNAs), we investigated whether our five identified smoking-associated sncRNA clusters represented full-length snoRNA transcripts or discrete processed fragments. To this end, we examined the distribution of sequencing reads across their corresponding canonical snoRNA sequences. We found that a single fragment predominated in every cluster, accounting for 24.0% (cl_193), 37.3% (cl_196), 60.8% (cl_201), 33.1% (cl_207), and 74.3% (cl_214) of the total cluster expression (Fig. 4, A, C, and Appendix B: Fig. S7, Table S11). When mapped onto the predicted secondary structures of their respective mature snoRNAs, these fragments spanned approximately half of the mature sequence, covering either the 5′ or the 3′ half (Fig. 4 B, D; Appendix B: Fig. S7).

**Fig. 4.**
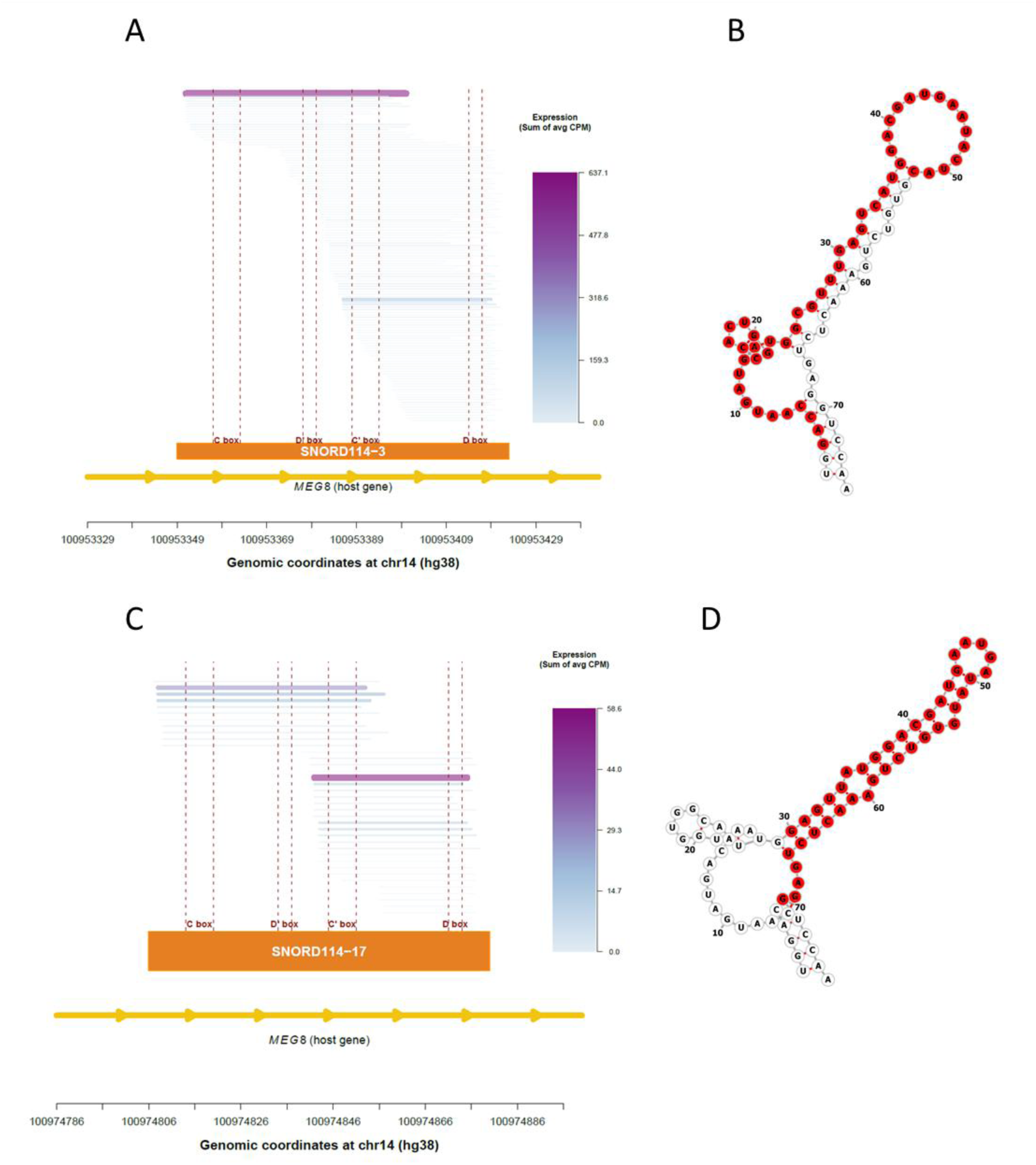
Characterization of the predominant sdRNAs detected in the significant smoking-associated clusters cl_201 (SNORD114-3 and piR-26394) and cl_207 (SNORD114-17). (A, C) Genomic distribution of the representative sequence families identified across the annotated SNORD114-3 (A) and SNORD114-17 (C) loci. Each horizontal segment represents a representative sequence family positioned according to its genomic coordinates. Sequence family expression (sum of average CPM) is indicated by both segment thickness and a color gradient ranging from white (low expression) to purple (high expression). The C, D, C′, and D′ box motifs are shown below each snoRNA. The fragment selected for downstream structural analyses is highlighted in purple. (B, D) Predicted minimum free energy secondary structures of SNORD114-3 (B) and SNORD114-17 (D). The corresponding sdRNA is highlighted in red. *CPM: counts per million; sdRNA: snoRNA-derived fragment*

## 4. Discussion

SncRNAs play important functions in gene regulation within the placenta^25^, an organ essential for fetal development. While some previous studies have characterized placental miRNAs^60–64^, far fewer have examined other sncRNA^19,26,28^ species and their relationship with environmental exposures ^17,29^. In this study, we applied the seqBuster/seqCluster pipeline to comprehensively characterize the placental sncRNA landscape, including miRNAs and other sncRNA classes, and investigated their associations with two prenatal environmental exposures, MSDP and air pollution, that may influence similar biological pathways. Both exposures have been associated with reduced birth weight and adverse health outcomes later in life^4,5^, as well as with molecular alterations in the placenta, including changes in DNA methylation^65,66^.

In the 390 placentas from the BiSC cohort, we identified 422 miRNAs and 979 sncRNA clusters, obtained using the seqBuster and seqCluster tools, respectively. Expression estimates of placental miRNAs obtained from both methods showed strong concordance, indicating robustness of the cluster-based approach. The TA50 metric revealed a highly skewed abundance distribution, with approximately 1% of detected features accounting for half of the total sncRNA abundance, suggesting that placental sncRNA expression is largely driven by a limited number of highly abundant features. Similar expression patterns have been described in the placenta and other tissues, where a restricted subset of highly abundant miRNAs contributes disproportionately to the overall small RNA landscape^67–69^.

We observed high overlap between the miRNA and snoRNA identities detected in our study and those reported by Gong *et al*. (2021), whereas piRNA annotations showed lower concordance across studies, including comparisons with Gong *et al*. (2021) and Martínez *et al*. (2021). Several of the most abundant miRNAs in our dataset, including miR-516, miR-126, miR-100 and miR-21, have previously been reported among dominant placental miRNAs^17,26,70^. Notably, miR-516, the most abundant miRNA detected in our study, belongs to the placenta-enriched C19MC^70^, highlighting the contribution of placenta-specific miRNA clusters to the placental small RNA landscape^14,16,17^. In contrast, placental profiles of snoRNAs and piRNAs showed few concordances with Gong *et al*. study, suggesting that these RNA classes may be more sensitive to differences in sequencing approaches, processing pipelines, and annotation strategies. Similarly, although tRFs have been associated with preeclampsia^71^, their expression patterns and biological functions in the placenta remain poorly characterized.

In our study, miR-421 was the only miRNA significantly associated with MSDP. Although no previous studies have linked placental miR-421 with MSDP, this miRNA has been reported to be downregulated in mouse lungs following cigarette smoke exposure^72^. In the placenta, miR-421 regulates *ZEB1*, a transcription factor involved in trophoblast invasion and migration^73^. Dysregulation of the miR-421/ZEB1 axis has been reported in preeclampsia^74^, suggesting that the downregulation observed in our study may reduce repression of *ZEB1* and thereby potentially enhance trophoblast invasion. As MSDP has been associated with lower risk of preeclampsia ^75^, a phenomenon known as the smoking paradox, the downregulation of miR-421 may represent a potential molecular mechanism underlying this association. As miR-421 is located within the PAR1 of chromosome X, we also evaluated potential sex differences in its expression; however, no significant differences were observed between female and male placentas. Apart from miR-421, previous studies have reported smoking-associated downregulation of placental miR-21, miR-16, and miR-146a^76^. Although these miRNAs were detected in our datasets, we were unable to replicate these findings.

Beyond miRNAs, the most striking finding of this study was the identification of five smoking-associated sncRNA clusters mapping to the *DLK1-DIO3* locus. This locus is one of the largest imprinted non-coding RNA domains in the human genome and is regulated by two imprinting control regions, which govern the transcription of paternally expressed protein-coding genes and a large maternally expressed polycistronic unit^77,78^. The maternal RNAs from this region have been shown to be essential for normal placental vascularization and nutrient transport in murine models^79^. Moreover, dysregulation of the *MEG8* lncRNA gene has been associated with trophoblast dysfunction and adverse pregnancy outcomes^80,81^.

The MSDP-associated clusters mapped to the *SNORD113* and *SNORD114* snoRNA families within the *MEG8* lncRNA gene. All the sncRNA clusters showed upregulation, suggesting that MSDP may influence regulatory mechanisms acting across the locus rather than affecting individual transcripts independently. Given the size range of the sequences mapping to the SNORD113 and SNORD114 snoRNAs, the detected sequences are likely sdRNAs rather than full-length transcripts. Within each MSDP-associated cluster, a single fragment accounted for 24-74% of the total abundance and originated from a defined region of the mature snoRNA. These findings support growing evidence that box C/D snoRNAs undergo regulated post-transcriptional processing to generate sdRNAs^82^ rather than representing products of random RNA degradation. Interestingly, the predominant fragment identified within several snoRNA clusters overlapped annotated piRNAs, supporting previous reports that box C/D snoRNAs can generate piRNA-like molecules through alternative processing^83^. These observations highlight the complexity of non-coding RNA annotation within the *DLK1-DIO3* locus and raise the possibility that MSDP influences the biogenesis or stability of these specific fragments, rather than solely affecting the expression of canonical full-length snoRNA transcripts.

To our knowledge, no previous studies have linked MSDP to SNORD113-, SNORD114-, or MEG8-derived piRNAs in the placenta. However, the *DLK1-DIO3* locus has been shown to be susceptible to environmental perturbations in murine models, including maternal diet, metabolic stress, and prenatal alcohol exposure, which have been associated with alterations in DNA methylation and non-coding RNA expression within this imprinted region^84,85^.

For air pollution exposure, no sncRNAs remained significant after multiple-testing correction. Previous studies on prenatal air pollution have primarily focused on placental DNA methylation and miRNA expression^29,86^, whereas the role of other sncRNA classes, including snoRNAs, piRNAs, and tRFs, remains poorly characterized. One study reported associations between PM_2.5 e_xposure and placental expression of miR-21, miR-146a, and miR-222^87^. However, none of these miRNAs were associated with PM_2.5 e_xposure in our dataset, even at the nominal significance level. This discrepancy may reflect differences in exposure assessment and methodology between the two studies. Among the nominally associated miRNAs in our study, we observed downregulation of two members of the let-7 family, miR-98 and let-7f-1-3p, in response to PM_2.5 a_nd NO_2,_ respectively. In silico downstream analysis identified *LIN28B* as a target of miR-98. *LIN28B* is an RNA-binding protein highly expressed in the placenta^26^, and is part of a well-established regulatory feedback loop with the let-7 family. Reduced *LIN28B* expression has been implicated in preeclampsia, potentially through impaired trophoblast invasion, defective syncytialization and increased inflammatory responses^88^. We also identified nominal downregulation of miR-19b-3p and miR-301b-3p following PM_2.5 e_xposure. Both miRNAs have previously been reported to be downregulated in placental tissue, particularly in female infants, following prenatal carbon monoxide exposure from household air pollution^29^.

Several factors may explain why air pollution-associated signals were less evident than those observed for MSDP. Unlike active smoking, which represents a relatively high-dose and direct exposure, ambient air pollution is more diffuse and heterogeneous, varying according to residential environment, daily activities, and lifestyle. Consequently, the effective dose reaching the placenta is likely lower and more variable, reducing the power to detect consistent molecular signatures. Moreover, the constituents of air pollution and tobacco smoke are different and hence have different toxicities, which could be translated into their differential effects on sncRNA landscape.

Our study had several notable strengths. First, we implemented a comprehensive bioinformatic pipeline combining seqBuster and seqCluster to characterize a broad spectrum of placental sncRNAs, moving beyond previous studies that have largely focused on miRNAs alone. Second, maternal exposure to air pollution was estimated using high-resolution hybrid models that incorporated both residential and workplace locations, thereby reducing exposure misclassification. Third, while MSDP was self-reported and inherently subject to potential misclassification, we nevertheless observed biologically consistent exposure-duration patterns and robust results across sensitivity analyses. Fourth, our study extended placental epigenetic research beyond DNA methylation by identifying exposure-associated changes in sncRNAs, highlighting post-transcriptional regulatory mechanisms that may more directly influence gene expression and placental function.

This study was subject to several limitations. First, the bioinformatic analysis of sncRNAs remains challenging because short reads frequently map to multiple genomic locations and annotation databases for non-miRNA sncRNAs remain incomplete. Although our combined seqBuster/seqCluster approach successfully captured a broader diversity of sncRNAs than either method alone, the ultimate interpretation of these features depends heavily on the current state of available annotations. Second, bulk small RNA sequencing captures the average expression profile across the tissue and lacks cell-type resolution. While we controlled for technical and biological variation in biopsy collection by adjusting for placental cell-type proportions estimated from DNA methylation data and including SVs to capture residual technical and biological variability, single-cell sequencing methods should be applied in future studies to resolve cell-type-specific effects. Third, our statistical power was constrained by the available sample size and sequencing depth, which may have limited our capacity to detect significant associations involving low-abundance sncRNAs. Fourth, functional interpretation remains speculative for many sncRNAs because experimental evidence and functional annotations for these molecules are still scarce. Finally, the predominantly European ancestry of the BiSC cohort may limit the generalizability of our findings to more diverse populations.

Overall, our findings provide a refined and novel characterization of the placental sncRNA landscape using a cluster-based approach capable of capturing diverse sncRNA species beyond miRNAs. We identified sncRNAs from the *DLK1-DIO3* imprinted locus, together with miR-421, as being significantly associated with MSDP. Although no associations with prenatal air pollution remained significant after multiple-testing correction, several suggestive candidate sncRNAs with potential roles in placental development and function were identified, warranting further investigation.

## Data Availability

Individual-level data may be available by contacting the BiSC cohort (https://projectebisc.org/en/bisc-project/publish-with-us/) and signing required approvals.

## Author contributions

**Álvaro Falcón-Gracia:** Writing – original draft, Writing – review and editing, Visualization, Methodology, Investigation, Formal analysis, Data curation, Conceptualization.

**Hana Vespalcová:** Writing – review & editing, Methodology, Data curation

**Dolors Peligrí:** Methodology, Data curation

**Sofia Aguilar-Lacasaña:** Data curation

**Lorena Pantano:** Methodology

**Ramil N Nurtdinov:** Methodology, Data curation

**Zoraida Garcia:** Data curation

**Alan Domínguez**: Writing – review and editing

**Laura Gómez-Herrera**: Data curation, Writing – review and editing

**Olga Sánchez**: Writing – review and editing

**Mireia Gascón**: Writing – review and editing

**Maria Foraster**: Writing – review and editing

**Roderic Guigó**: Writing – review and editing

**Elisa Llurba**: Writing – review and editing

**Maria Dolores Gomez-Roig**: Writing – review and editing

**Ioar Rivas**: Writing – review and editing

**Payam Dadvand:** Writing – review and editing, Supervision, Investigation, Funding acquisition, Conceptualization.

**Jordi Sunyer:** Writing – review and editing, Supervision, Investigation, Funding acquisition, Conceptualization.

**Mariona Bustamante:** Writing – review and editing, Supervision, Data Curation, Methodology, Investigation, Conceptualization.

**Marta Cosin-Tomas:** Writing – review and editing, Supervision, Data Curation, Methodology, Investigation, Conceptualization, Funding acquisition.

## Ethics

From pregnancy up to 18 months’ visits were approved by the Clinical Research Ethics Committee of the Parc de Salut Mar project (2018/8050/I), Medical Research Committee of the Fundació de Gestió Sanitària del Hospital de la Santa Creu i Sant Pau de Barcelona (EC/18/206/5272), and Ethics Committee of the Fundació Sant Joan de Déu (PIC-27-18). Before joining the cohort during their regular first trimester hospital visit, participants were informed by a BiSC midwife or nurse about the study’s details, duration, and their option to withdraw without penalty. If they agreed to take part, they signed consent forms permitting the collection of biological samples and genetic studies, receiving a copy for themselves. Parents or legal guardians of children also provided informed consent before any study procedures commenced. Specific consent forms for genetic studies were also provided to mothers and parents or legal guardians of children.

## Acknowledgments

We would like to thank all the participants and their families for their generous collaboration. A full list of BiSC researchers can be found (https://projectebisc.org/en/team/)

The authors acknowledge the use of ChatGPT (GPT-5, OpenAI, San Francisco, CA, USA) to improve the clarity and readability of the manuscript. The authors reviewed and edited the content as necessary and take full responsibility for the final text.

## Abbreviations

BC: blackcarbon
BiSC: Barcelona Life Study Cohort
BMI: body mass index
C14MC: chromosome 14 miRNA cluster
C19MC: chromosome 19 miRNA cluster
CPM: counts per million
DMR: differentially methylated region
DOHaD: Developmental Originsof Health and Disease
ENT: effective number of independent tests
GTEx: Genotype-Tissue Expression
HPA: human protein atlas
HSjD: hospital Sant Joan de Déu
HSP: hospital Sant Pau i la SantaCreu
IQR: inter quartile range
lncRNA: long non-coding RNA
Log2FC: log2 fold change
LUR model: land use regression model
M: million
miRNA: microRNA
MSDP: maternal smoking duringpregnancy
NO2: nitrogen dioxide
nRBCs: nucleated red blood cells
nt: nucleotide
PAR1: pseudoautosomal region 1
PCA: principal component analysis
piRNA: piwi-interacting RNA
PM: particulate matter
PM2.5: particulate matter with diameter ≤ 2.5 μm
POPs: Pregnancy Outcome Prediction study
QC: quality control
RIN: RNA integrity number
rRNA: ribosomal RNA
sdRNAs: snoRNA-derived fragments
SES: socioeconomic status
SMD: standarized mean difference
sncRNA: small non-coding RNA
snoRNA: small nucleolar RNA
snRNA: small nuclear RNA
SV: surrogate variable
SVA: surrogate variable analysis
TA50: transcriptome abundance 50
TMM: trimmed mean of the m values
TPM: transcript per million
tRF: tRNA-derived fragment
tRNA: transfer RNA
UTR: untranslated region

